# Analysis of a paediatric cohort of dyslipidaemic patients using unsupervised learning methods provides insights into the biochemical phenotypes of familial hypercholesterolemia

**DOI:** 10.1101/2022.07.17.22277724

**Authors:** Marta Correia, Mafalda Bourbon, Margarida Gama-Carvalho

## Abstract

Familial hypercholesterolaemia (FH) is a highly prevalent silent disease with known genetic causes and poor prognosis if undiagnosed into adulthood. Characterised by high levels of total cholesterol and low-density lipoprotein cholesterol from birth, the majority of cases that fit the clinical criteria for FH do not present mutations in the disease associated genes and seem to result from polygenic and/or environmental causes. In this study we have addressed the heterogeneity of extended blood biochemical and genetic parameters across a cohort of children using an unsupervised hierarchical clustering approach. In addition to correctly classifying individuals into the two classes recognized in clinical studies for familial hypercholesterolaemia (with and without genetic diagnosis), a subset of patients with mixed characteristics was systematically identified as representing a third category. The careful analysis of biochemical, genetic, and anthropomorphic characteristics that constitute hallmarks of each group provides detailed insights into the characteristics of each group, contributing to unravel the complexity of FH and dyslipidaemic phenotypes. The results presented here may assist in the future identification of novel biomarkers to efficiently identify FH+ individuals.

## INTRODUCTION

Familial hypercholesterolaemia (FH), an autosomal co-dominant disorder, is the most common monogenic dyslipidaemia, with an estimated prevalence of 1/250 for heterozygotes worldwide [1], [2]. The prevalence of homozygous FH is approximately 1/300 000. This condition usually results from loss-of-function genetic variants in the *LDLR* gene and, less commonly, in the *APOB* gene. Gain-of-function variants in *PCSK9* gene also produce a similar phenotype, although it is a rare cause of FH [1], [3].

FH is clinically characterised by high levels of low-density lipoprotein cholesterol (LDL-C) since birth and consequent accumulation of cholesterol within blood vessels, which triggers the development of atherosclerosis and premature cardiovascular disease (i.e., at 20-39 years old). FH is still widely under-diagnosed and undertreated, including in Portugal, where no prior clinical or genetic studies were available before the beginning of the Portuguese FH study (PFHS) in 1999 [4], [5]. An early diagnosis of FH is essential to improve patient prognosis through appropriate therapeutic measures, genetic counselling, and access to specialised medical services [5], [6].

Given the silent nature and prevalence of FH, current guidelines support the testing of the disease hallmark genes (*LDLR, APOB* and *PCSK9*) in patients that comply with the clinical diagnostic criteria, and cascade screening of their family members [7]. However, most hyperlipidaemic subjects do not have a monogenic defect [1], [8]. Rather, their disease is most likely established through a polygenic genetic background, with a variable environmental contribution modulating the phenotypic expression [1], [8]. Although the lipid profile of polygenic subjects is usually less severe than that of FH subjects regarding total cholesterol (TC) and LDL-C levels, the differences are often subtle enough to prevent an accurate distinction between the two [9]. As a consequence, the yield of FH genetic screening programs is relatively low, assuming significant costs for patients and/or national health systems. Thus, new approaches are needed to achieve a better distinction between monogenic and polygenic dyslipidaemia patients, preferably based on methods that support a more accurate and less time-consuming diagnosis, with an acceptable cost/benefit ratio. Application of integrative data analysis tools, like machine learning (ML)-based methods, is expected to assist in the identification of sub-groups of individuals characterised by specific biological parameters, thus supporting the identification of novel biomarkers that can best differentiate FH patients [10], [11].

ML involves the development of statistical models and algorithms that can progressively learn from data and achieve desired performance on a specific task, especially when it is not possible to manually develop a set of rules based on all the intrinsic characteristics of the data. Comparatively to traditional statistical methods, ML has better performance at finding patterns within complex datasets like, for example, clinical data. Therefore, the application of ML based-methods can contribute to the identification of reliable disease biomarkers and improve patient stratification systems [11]. Within ML, we can find both supervised and unsupervised learning methods. In our previous study [12], a supervised ML approach was used to develop new classification models able to improve the selection of dyslipidaemic individuals for FH genetic testing. The current work followed an unsupervised ML approach applied to the same dataset, which comprises the clinical, genetic and extensive blood lipid profile of a sample of 211 children (from 2 to 17 years old) from the Portuguese FH study (PFHS), which we designated PFHS-ped. Unlike most adult subjects in the PFHS study, this cohort was not under any medication, thus ensuring their lipid profile reflects a “natural” scenario. Furthermore, given the global aim of identifying FH patients as early as possible, we chose to focus our study on a paediatric cohort to avoid additional confounding factors.

Unsupervised learning aims to identify patterns within an unlabelled input dataset, for example through the application of methods based in dimensionality reduction (e.g., principal components analysis, PCA) or clustering analysis for identification of different groups/clusters of subjects within the dataset [11]. The method used in this work, hierarchical clustering, organises data into a hierarchical structure based on appropriate (dis)similarity or distance measures between every pair of subjects in the dataset. In the case of agglomerative nesting, the clustering algorithm groups data by means of a sequence of partitions starting with each unit (subject) forming a separate cluster and then merging similar clusters into larger clusters [13]–[15]. The results of these approaches can thus support a better understanding of the degree of heterogeneity that can be found in a given population/dataset. In this work, we addressed this issue within the PFHS-ped cohort, with the final aim of guiding the development of better classification rules for young patients presenting with clinical features of FH.

## METHODS

### The PFHS-ped dataset: patient selection, biochemical and clinical data

The work dataset – PFHS-ped – has been previously described [12]. In summary, it comprises a cohort of 211 children (from 2 to 17 years old) not undergoing statin treatment at the time of referral and for which body mass index (BMI), genetic testing data and a basic set of lipid parameters were available [12]. 68% of the 211 individuals in PFHS-ped fulfil the Simon Broome (SB) clinical criteria for FH [18], while the rest present TC above the 95^th^ percentile for their age and sex and a family history of hypercholesterolaemia [17]. The PFHS-ped data comprises the extended characterization of blood lipid parameters described in Table 1, divided into three profiles: “Basic”, “Advanced” and “Lipoprint”, for commonly determined, specialized and Lipoprint test lipid parameters, respectively, and is available as supplementary material to reference [12].

**Table 1.** Description of the biochemical parameters and ratios in each lipid profile – “Basic”, “Advanced” and “Lipoprint”. N/A: not applicable

| Profile | Parameters |  | Units | Description |
| --- | --- | --- | --- | --- |
| Basic | Biochemical | TC | mg/dl | total cholesterol |
|  |  | LDL-C |  | low-density lipoprotein cholesterol |
|  |  | HDL-C |  | high-density lipoprotein cholesterol |
|  |  | TG |  | triglycerides |
|  |  | Lp(a) |  | Lipoprotein (a) |
|  |  | ApoB |  | Apolipoprotein B |
|  |  | ApoA-I |  | Apolipoprotein A-I |
|  | Rati | ApoB/ApoA-I | N/A | pro-atherogenic vs anti-atherogenic ratio |
|  |  | TG/ApoB |  | TG metabolism vs LDL metabolism ratio |
|  |  | TC/HDL-C |  | pro-atherogenic vs anti-atherogenic ratio |
| Advanced | Biochemical | ApoA-II | mg/dl | Apolipoprotein A-II |
|  |  | ApoC-II |  | Apolipoprotein C-II |
|  |  | ApoC-III |  | Apolipoprotein C-III |
|  |  | ApoE |  | Apolipoprotein E |
|  |  | sdLDL.Day |  | Small dense LDL |
|  | Ratios | ApoC-II/ApoC-III | N/A | anti-atherogenic vs pro-atherogenic ratio |
|  |  | sdLDL/LDL-C |  | most atherogenic LDL in total LDL-C |
| Lipoprint | Biochemical | VLDL | mg/dl | Very low-density lipoprotein |
|  |  | MIDA |  | IDL fraction A |
|  |  | MIDB |  | IDL fraction B |
|  |  | MIDC |  | IDL fraction C |
|  |  | LDL1 |  | Buoyant (large) LDL fraction 1 |
|  |  | LDL2 |  | Buoyant (large) LDL fraction 2 |
|  |  | HDL.Lipo |  | High-density lipoprotein |
|  |  | sdLDL.Lipo |  | Small dense LDL (fractions 3 to 7) |
|  |  | IDL |  | Intermediate-density lipoprotein |
|  | Ratios | VLDL/IDL | N/A | TG metabolism vs LDL metabolism ratio |
|  |  | VLDL/LDL-C |  | TG metabolism vs LDL metabolism ratio |

In addition to quantitative variables (BMI, age, and the biochemical parameters present in Table 1), PFHS-ped comprises a set of categorical variables that were used as supplementary variables: “Class”, classification based on FH genotype (FH+ or FH-); “Gender” (female or male); “Activity class”, according to the percentage of molecular activity that is kept by the affected gene allele; “Gene”, the affected gene in FH+ individuals (*LDLR, APOB, PCSK9*); “SB criteria”, concerning the fulfilment of TC and/or LDL-C cut-offs of the SB clinical criteria (yes or no); “Lipoprint profile”, according to low or high concentration of sdLDL in serum as measured by the Lipoprint assay (profile A or B, respectively); and “LDL-C score”, a polygenic risk score associated with LDL-C levels and based on a panel of six SNPs, as performed by Mariano et al. [23]. The categories of “LDL-C score”, “Activity class” and the variable “BMI class” were established for the purpose of this study. The variable “LDL-C score” comprises four categories that correspond to the following, from high to low polygenic contribution: ≥0.9, 0.9-0.7, 0.7-0.5, <0.5. Regarding “Activity class”, variants were divided in the following categories: “null variants”, presenting less than 2% of activity compared to wild type allele; “defective variants” with different degrees of molecular activity corresponding to three categories (2-20%, 20-40%, 40-65%); “null_pred variants”, predicted to be null according to *in silico* analysis; “def_pred variants”, predicted to be defective according to *in silico* analysis. Concerning variable “BMI class”, it comprises the classification of BMI according to gender and age, following the percentiles for children and adolescents from the World Health Organization (WHO) [24], [25]. Accordingly, “BMI class” includes the following categories: severe thinness, thinness, normal, overweight, obesity.

### Hierarchical clustering and statistical analysis

All data analysis was performed using R software (R version 3.4.3) [26]. For the hierarchical clustering of principal components (HCPC) analysis, we used *PCA* and *HCPC* functions of the package *FactomineR* (version 1.41) [27].

The *PCA* function performs PCA with the possibility of adding supplementary individuals and variables, both quantitative and categorical. This supplementary data does not contribute to PCA itself but can be useful for results interpretation [28]. In the current data analysis, “Class” was used as a discriminant factor while observing the distribution of individuals classified as FH+ or FH- across the clusters. For the “All” subset, since it was the subset that showed the most well-defined cluster partition (see results), other categorical variables besides “Class” were included as supplementary data to test their association with clusters, such as “BMI class”, “Activity class”, “Gender”, “Gene”, “SB criteria”, “Lipoprint profile”, and “LDL-C score”.

The *HCPC* function performs an agglomerative hierarchical clustering on the results of PCA, by using the PCA coordinates of individuals to measure the distance between them. For this, HCPC uses Ward’s minimum variance method that aggregates clusters when it translates in a minimum of within-variance growth, thus contributing for homogenous clusters [29]. In this study, the hierarchical clustering was performed using the first five dimensions of PCA (default option), which explain most of the data variance. The obtained hierarchical tree, also known as dendrogram, was cut at the suggested level, corresponding to a partition in three clusters. HCPC output also includes a description of the clusters by individuals, variables, and dimensions; assignment of a cluster for each patient; other graphic visualisations besides the dendrogram, such as cluster maps [30]. The functions *fviz_dend* and *fviz_cluster* of the package *factoextra* were applied to improve the visualisation and interpretation of dendrograms and cluster maps [31]. Both functions allow an enhanced ggplot2-based visualisation of the plots [32].

For the characterization of clusters by quantitative variables, the correlation ratio was directly measured by the HCPC algorithm between each variable and the cluster partition (i.e., cluster assignment of individuals, called “cluster variable” by HCPC). This was followed by a student’s t-test to determine the variables whose correlation ratio was significantly different from zero, and thus have a significant contribution for cluster partition. In addition, HCPC measures the average of a variable in the cluster (named as “mean in category”) and in the whole subset (named as “overall mean”), including the associated standard deviations. The HCPC algorithm applies a student’s t-test to enquire, for each variable, if the difference between the mean in category and the overall mean was statistically significant under the confidence level of 95%. A positive or negative value of the test statistics indicates if the mean in category is greater or lower than the overall mean, respectively.

Regarding categorical variables, a chi-squared test was performed to assess the association between each variable and the cluster partition. Then, for each variable category, a statistical test was performed to check if the proportion of individuals within the category that belong to a given cluster was significantly different from the proportion of individuals assigned to the same cluster that belong to this category. This allowed us to assess which categories were underrepresented or overrepresented in each cluster. For a description of clusters by individuals, the distance between each individual coordinate and the gravity centre of the assigned cluster was measured to assess the paragons, or individuals that most characterise each cluster because they are the closest to the cluster centre. In addition, measuring the distance between each individual coordinate and the gravity centre of other clusters allowed the assessment of the most specific individuals of each cluster, which correspond to the farthest individuals to the centre of the other clusters. For cluster characterization by dimensions, an identical analysis to that carried out for quantitative variables was performed, considering each PCA dimension a quantitative variable composed by individual coordinates. The methodology behind HCPC analysis is explained in more detail by Husson et al. [29] and Lê et al. [27].

For prediction of class probabilities of individuals from the “All” subset, the best trained model from our previous study (Imp_B model) [12] was applied in that subset. The basic function *predict* was used with the argument “type” set for probabilities. Then, each of the 78 individuals was assigned to a predicted class (FH+ or FH-) and to the probability of belonging to each of the two classes. The sum of both probability values was equal to 1. In addition, the difference between probabilities (called Δprob) was measured for each individual, using absolute values of probability. Categories were established for Δprob taking into account that the lower the value, the more ambiguous is the classification of individuals.

For all the figures showing dendrograms, *dplyr* [33] and *ggplot2* [34] packages were used to assign a different colour to each variable category and plot the variables aligned with individuals by their order of appearance in the dendrogram.

## RESULTS

### Clustering analysis of the PFHS-ped cohort consistently describes three hierarchical groups

To unravel the biochemical patterns among FH+ and FH- individuals, while searching for potential biomarkers that may improve patient discrimination, a hierarchical clustering of principal components (HCPC) analysis was carried out separately on seven distinct subset of the PFHS-ped. These data subsets were previously defined to overcome the lack of a complete lipid profile for all the 211 individuals that comprise the PFHS-ped dataset, allowing us to make the best possible trade-off for group size (ranging from 78 to 211 individuals) and available data (all possible combinations of the three lipid profiles – Basic, advanced and Lipoprint, as described in methods) [12]. Given our aim to perform an unsupervised classification of the PFHS-ped cohort regarding FH status, the classification of individuals as FH+/FH- (variable “Class”) was only considered for interpretation of clustering results and thus did not contribute to the establishment of clusters.

Unexpectedly, the results of the clustering approach showed that in all subsets individuals were distributed in three distinct clusters. This observation is in stark contrast to the traditional assignment of patients into two classes based on their genetic profile (FH+ and FH-). This suggests the existence of a third group of individuals characterised by a distinct lipidic pattern, based on a diverse set of biochemical parameters. To obtain insights into the nature of these three groups, we looked at the distribution of FH+ and FH- individuals among clusters. As shown in Figure 1, in every subset it was possible to identify a pattern in the distribution of individuals according to their class. One of the clusters was always predominantly composed of FH+ patients and another cluster displayed a predominance of FH- individuals. The third cluster consistently presented a “mixed” population including a considerable number of both FH+ and FH- individuals. In spite of the smaller size when compared to the other data subsets, the “All” subset presented the best defined distribution of individuals among the three clusters, regarding FH+/FH- classification (Figure 1). After the “All” subset, the best distribution pattern was found in the subsets “Basic & Advanced”, “Basic & Lipoprint” and “Advanced & Lipoprint”. This suggests that a combination of parameters from different lipid profiles contributes to a better distinction between individuals, which is in agreement with previous results obtained using a supervised ML approach considering a population with two classes [12].

**Figure 1.**
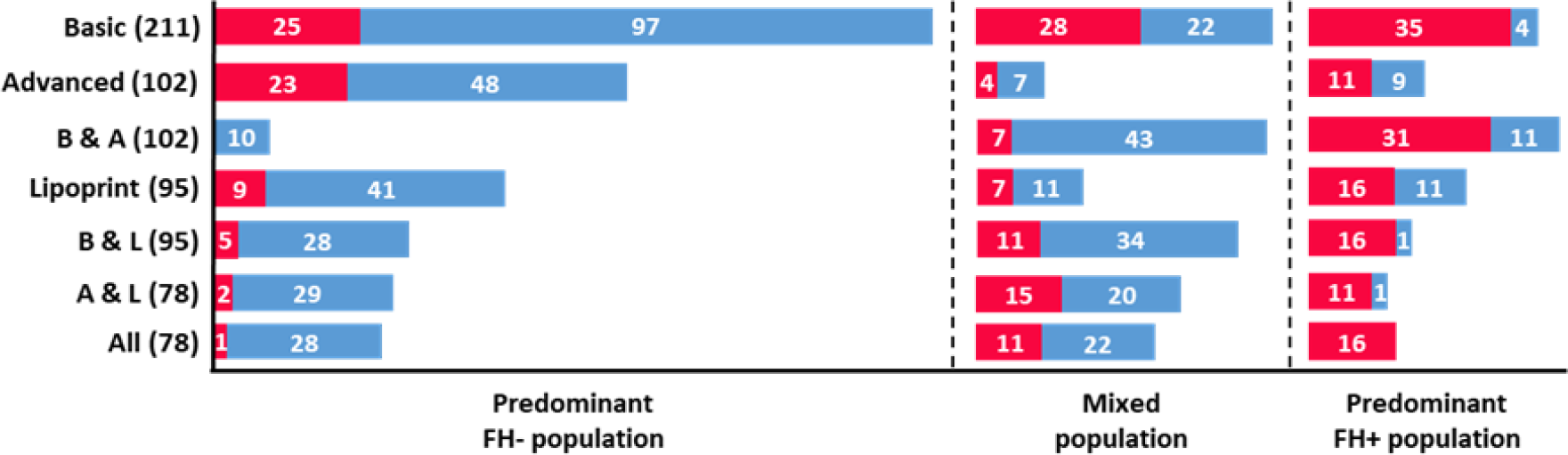
Characterization of clusters regarding the number of individuals according to their class, for each of the seven subsets. Red and blue bars correspond to FH+ and FH- populations, respectively. The number of individuals for each class is present in white. Each subset presents the total number of individuals within brackets. B & A: “Basic & Advanced”; B & L: “Basic & Lipoprint”; A & L: “Advanced & Lipoprint”.

Given this observation, a detailed exploration of the parameters behind clustering was focused on the “All” subset (called from this point on “working population”), whose distribution of individuals by clusters is shown in detail in Figure 2. It is noteworthy that the cluster with the so called “mixed population” is split into two branches, which are enriched in either FH+ or FH- individuals.

**Figure 2.**
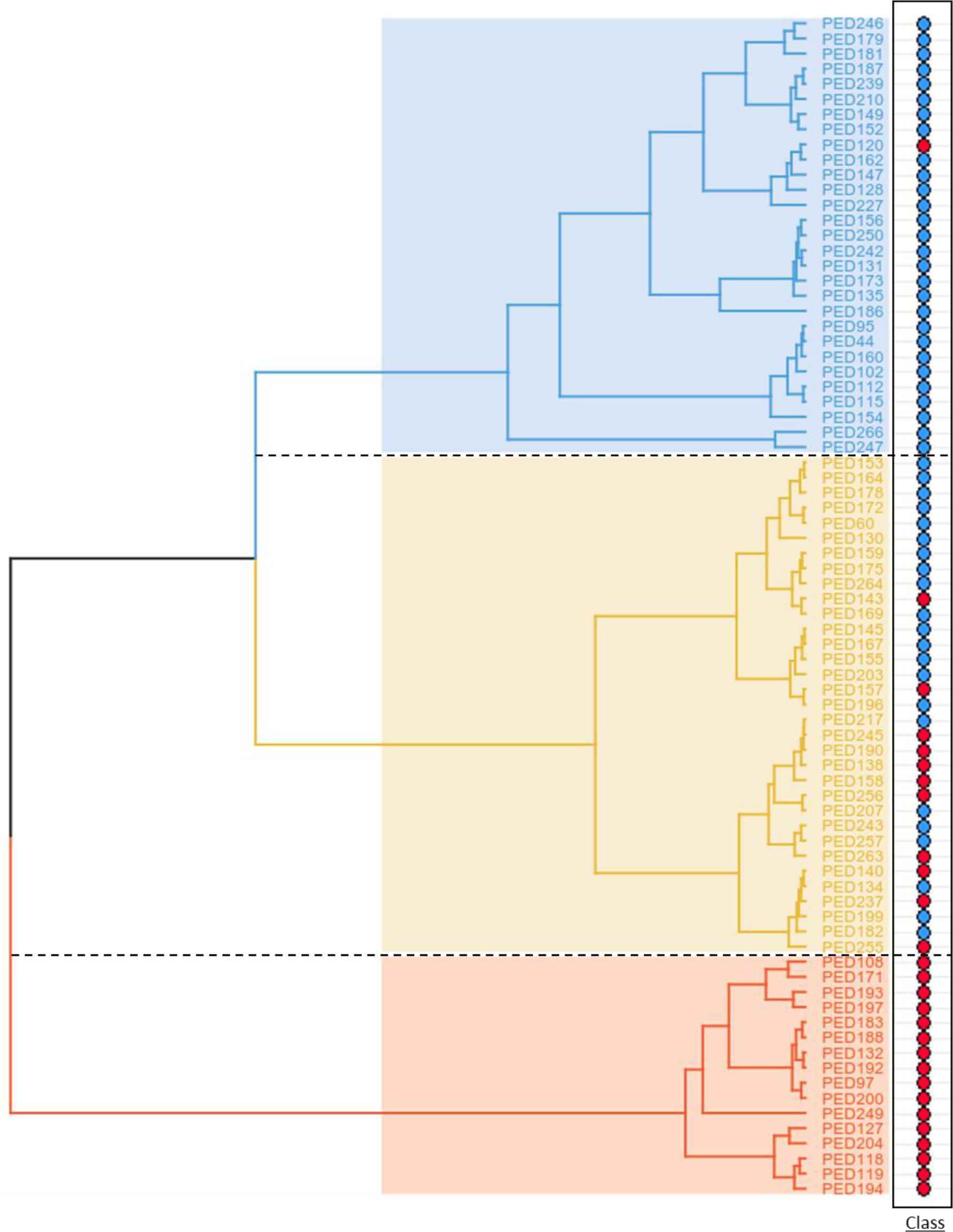
Dendrogram of the “All” subset showing the distribution of individuals in three clusters. Blue, yellow and red coloured clusters correspond to predominant FH-, mixed, and predominant FH+ populations, respectively. In the right panel, red and blue circles identify FH+ and FH- individuals, respectively.

### Cluster description by quantitative variables identifies metabolic pathways of interest for differentiation of FH+ and FH- classes

As mentioned before (see methods), the HCPC analysis allows the acquisition of a detailed description of clusters regarding the contribution of both quantitative and categorical variables, individuals, and dimensions. This information is important for an accurate characterization of clusters and to explain the distribution of individuals within each cluster, as well as to identify potential lipid profile patterns. To understand which parameters contributed to the cluster partition of the working population, the statistical analysis carried out by HCPC was carefully explored. This analysis considered all the quantitative variables used by the clustering algorithm, which comprise all the biochemical parameters from PFHS-ped, BMI and age. Accordingly, the results included a ranked list of the statistically significant parameters for the distribution of individuals among three clusters (Table 2). The parameters were ranked according to the correlation ratio between each parameter and cluster partition, which means that parameters with the highest correlation are at the top of this list. TG/ApoB was the parameter that mostly contributed to the cluster partition, followed by VLDL, LDL-C, apoB/apoA-I and TC/HDL-C. Conversely, MIDA, HDL-C, Lp(a), VLDL/IDL and sdLDL/LDL-C were the five parameters with the smallest significant contribution to the clustering. From a total of 30 quantitative variables, only five were not considered as statistically significant for the establishment of clusters (i.e., the correlation ratio was not significantly different from zero): apoC-II/apoC-III, apoE, LDL2, and Lipoprint measurements of HDL and sdLDL.

**Table 2.** Ranked list of the statistically significant parameters for the distribution of individuals by three clusters, under the level of significance of 95%. The parameters were ranked from the highest to the lowest significant contribution to cluster partition, according to the correlation ratio (**η**^**2**^) between each parameter and cluster division.

| **Rank** | **Variables** | **$\eta^2$** | **p-value** |
| --- | --- | --- | --- |
| 1 | TG/ApoB | 0.56 | 3.07E-14 |
| 2 | VLDL | 0.55 | 9.93E-13 |
| 3 | LDL-C | 0.53 | 2.58E-12 |
| 4 | ApoB/ApoA-I | 0.51 | 7.49E-12 |
| 5 | TC/HDL-C | 0.45 | 2.21E-11 |
| 6 | IDL | 0.43 | 1.61E-10 |
| 7 | VLDL/LDL-C | 0.43 | 1.32E-09 |
| 8 | ApoC-III | 0.43 | 1.45E-09 |
| 9 | MIDB | 0.42 | 2.71E-08 |
| 10 | TC | 0.41 | 3.62E-08 |
| 11 | ApoB | 0.39 | 8.06E-08 |
| 12 | LDL1 | 0.35 | 1.95E-07 |
| 13 | TG | 0.34 | 4.31E-07 |
| 14 | MIDC | 0.26 | 1.77E-06 |
| 15 | BMI | 0.24 | 7.53E-06 |
| 16 | ApoA-I | 0.23 | 1.46E-05 |
| 17 | ApoA-II | 0.23 | 6.33E-05 |
| 18 | sdLDL.Day | 0.21 | 7.49E-05 |
| 19 | ApoC-II | 0.18 | 3.08E-04 |
| 20 | Age | 0.18 | 6.93E-04 |
| 21 | MIDA | 0.17 | 1.03E-03 |
| 22 | HDL-C | 0.17 | 1.91E-03 |
| 23 | Lp(a) | 0.14 | 4.06E-03 |
| 24 | VLDL/IDL | 0.13 | 9.73E-03 |
| 25 | sdLDL/LDL-C | 0.09 | 1.53E-02 |

Once again, the importance of combining parameters from different lipid profiles to identify consistent lipid patterns among individuals is shown by the presence of “Basic”, “Advanced” and “Lipoprint” parameters (Table 2). Another important aspect is the presence of several ratios as significant contributors to cluster partition, which highlights the relationships between different parameters in the context of lipid metabolism.

For a better characterization of each cluster and to look for potential biochemical patterns, we explored the result of student’s t-test comparing the mean of each quantitative variable in the total working population of 78 individuals (i.e., “overall mean”) with the mean of the same variable in each cluster (i.e., “mean in category”). Figure 3 shows the list of parameters whose “mean in category” was significantly different from the “overall mean”. These are the parameters that best characterise each cluster.

**Figure 3.**
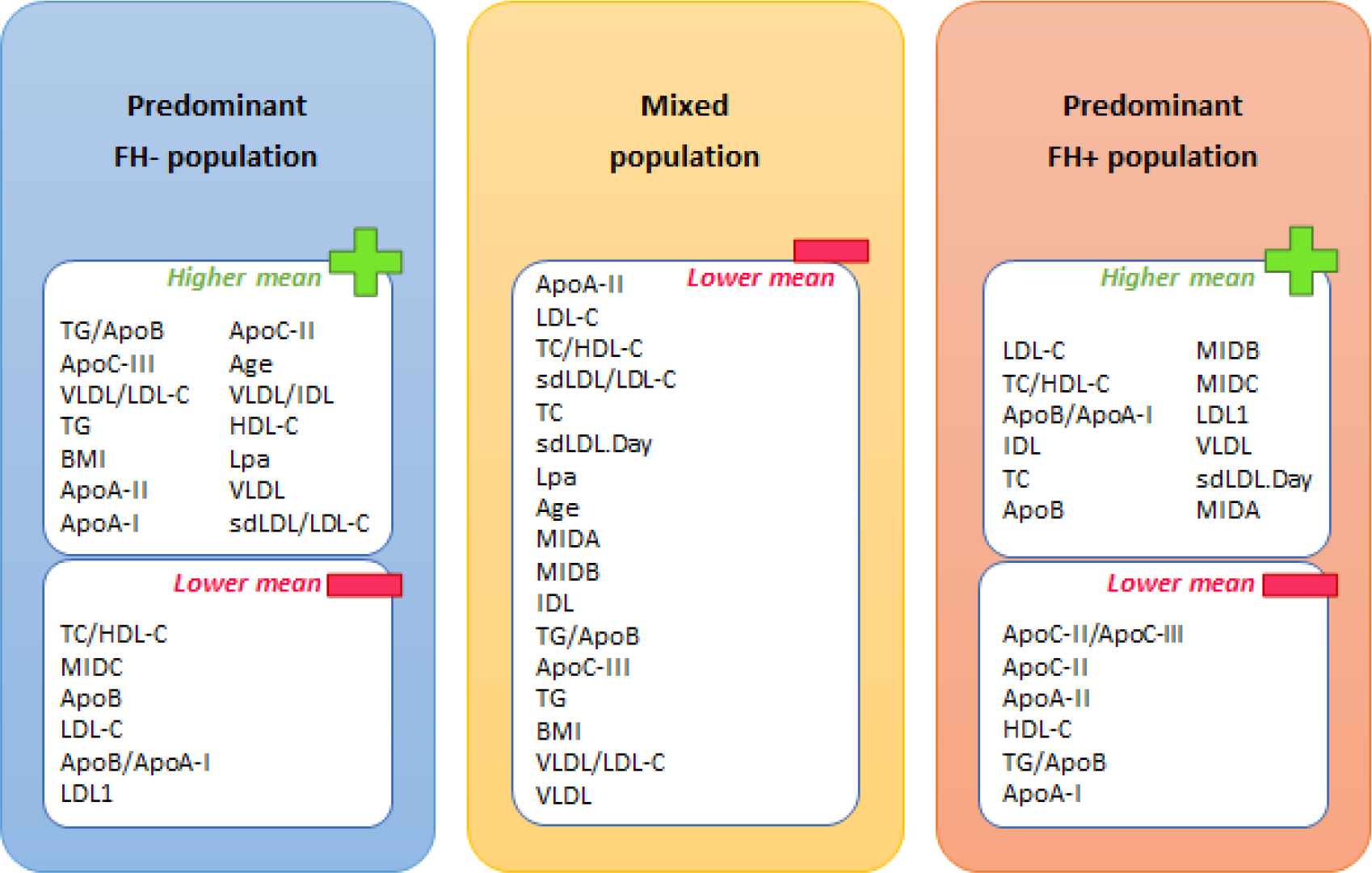
Parameters that best characterise each cluster, according to the difference between the “mean in category” of a given parameter and its “overall mean”. Among the significant parameters, those that present higher/lower mean values in the cluster in comparison to the overall mean are represented in separate boxes within the cluster. The level of significance of 95% was considered for this analysis.

The cluster with a predominant FH- population is mainly characterised by higher values of parameters related to TG metabolism and lower values of parameters related to LDL and apoB metabolism. The inverse association was found in the cluster with FH+ as the predominant population. Regarding the mixed population, both TG and LDL/apoB related parameters have lower mean values than in the working population. Further details of each parameter distribution and mean trend among clusters are presented in Supplementary Figure 1.

### Cluster description by categorical variables identifies molecular, biochemical, and anthropomorphic patterns among individuals

Concerning the characterization of clustering results by a set of categorical variables associated with PFHS-ped (Supplementary Table 1), a chi-squared test was used to measure the association between each categorical variable and cluster partition. Table 3 presents the variables that significantly explain the distribution of individuals by three clusters. These variables did not contribute to cluster partition, instead they were included in clustering analysis as supplementary variables and thus only used for interpretation purposes.

**Table 3.** Categorical variables that present a statistically significant association with cluster partition results, under the confidence level of 95%.

| **Variables** | **p-value** |
| --- | --- |
| Class | 7.83E-10 |
| Gene | 3.52E-08 |
| Activity class | 1.53E-06 |
| SB criteria | 3.54E-03 |
| BMI class | 1.75E-02 |

As previously shown by Figure 2, the classification of individuals as FH+/FH- is closely associated with their distribution among clusters (Table 3). The affected gene in FH+ patients (*LDLR, APOB, PCSK9*) is also associated with clustering results, as well as the percentage of molecular activity that is kept by the affected allele (variable “Activity class”). These results show the relevance of genotype to patients differentiation. In addition, the fulfilment of SB criteria and BMI class are also significantly associated with cluster distribution, which suggest the presence of different degrees of disease severity among individuals and the potential contribution of environmental factors (e.g., rich fat diet) for the establishment of cluster patterns.

For a more detailed description of each cluster, the association between each of them and categorical variables was tested. Figure 4 presents the categories of different variables that showed a statistically significant association (positive or negative) with each cluster. Of note, none of the categorical variables presented a significant association with the mixed population cluster.

**Figure 4.**
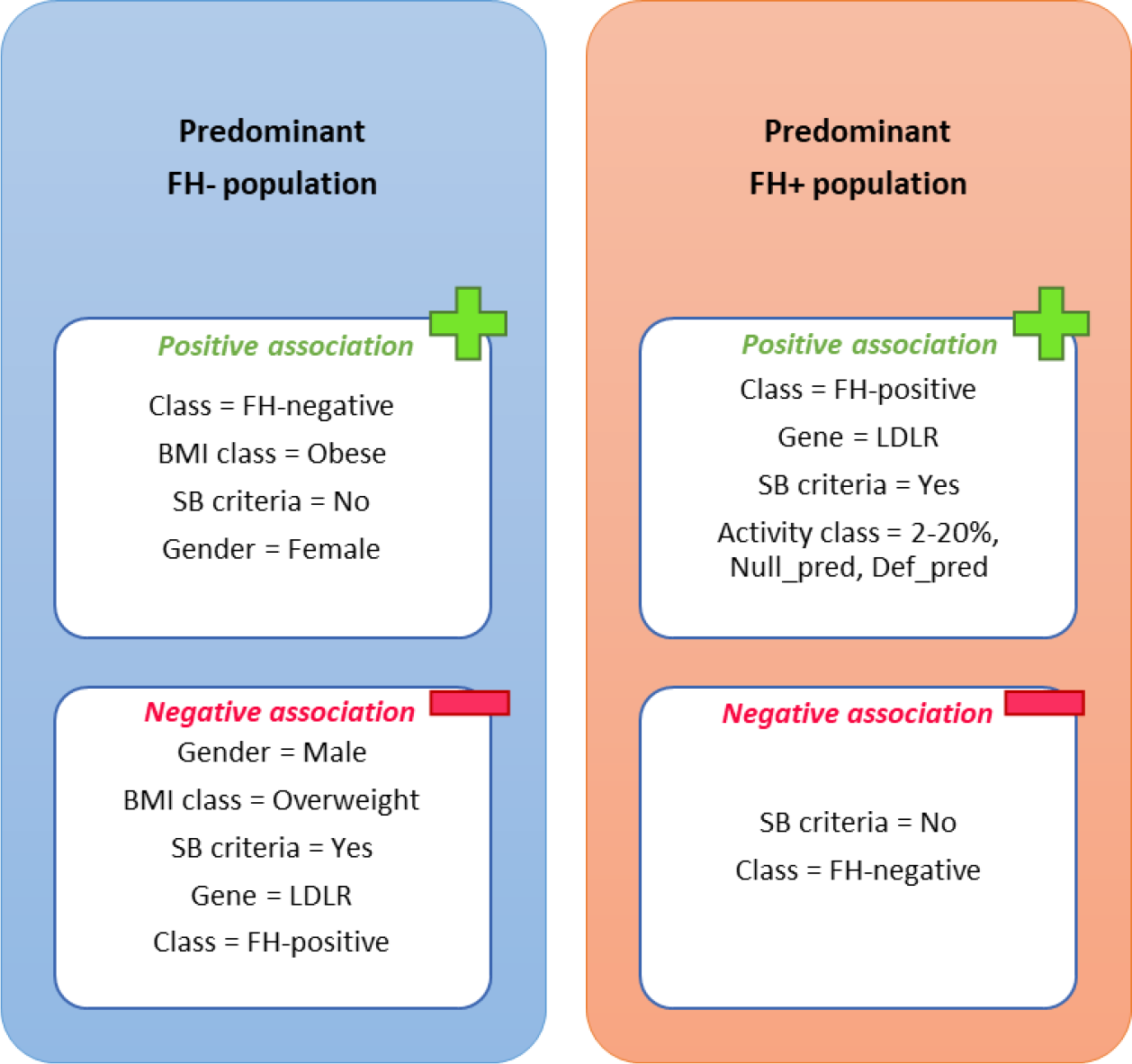
Variables categories that present a statistically significant association, positive or negative, with each of the clusters. Mixed population is not represented since no significant association was found between this cluster and categorical variables. The level of significance of 95% was considered for this analysis. The categories ‘Null_pred’ and ‘Def_pred’ of activity class are related to the results of *in silico* predictions for allele activity, corresponding to predicted null and defective variants, respectively.

As seen above, there is a clear pattern in the distribution of individuals among clusters according to their classification as FH+ or FH-. The variable “SB criteria”, regarding the fulfilment of TC and LDL-C cut-offs from the SB clinical criteria, also showed a significant association with clusters. Accordingly, the predominant FH+ cluster was associated with higher levels of TC and LDL-C, compared to the predominant FH- cluster that appeared to present a milder biochemical profile. Giving the fact that *LDLR* is the most affected gene in FH [9], the presence of a positive association between the predominant FH+ cluster and an affected *LDLR* gene, as well as the negative association of *LDLR* with the predominant FH- cluster is not surprising. In addition, the predominant FH+ cluster seemed to be characterised by a low activity of the affected allele (2-20% category), in addition to being associated with null and defective *in silico* predicted variants (“Null_pred” and “Def_pred” variants, respectively). This suggests that the predominant FH+ cluster is associated with a more severe genotype, with the significant presence of variants keeping a molecular activity not higher than 20% in comparison to wild type. Regarding other variables like gender and BMI class, the predominant FH- cluster appeared to be characterized by the presence of obese girls, in contrast to the negative association with overweight boys. Still, this association of individual distribution among clusters with gender should be considered with caution, as explained below.

Figure 5 attempts to provide a clear visualisation of cluster characterization by categorical variables.

**Figure 5.**
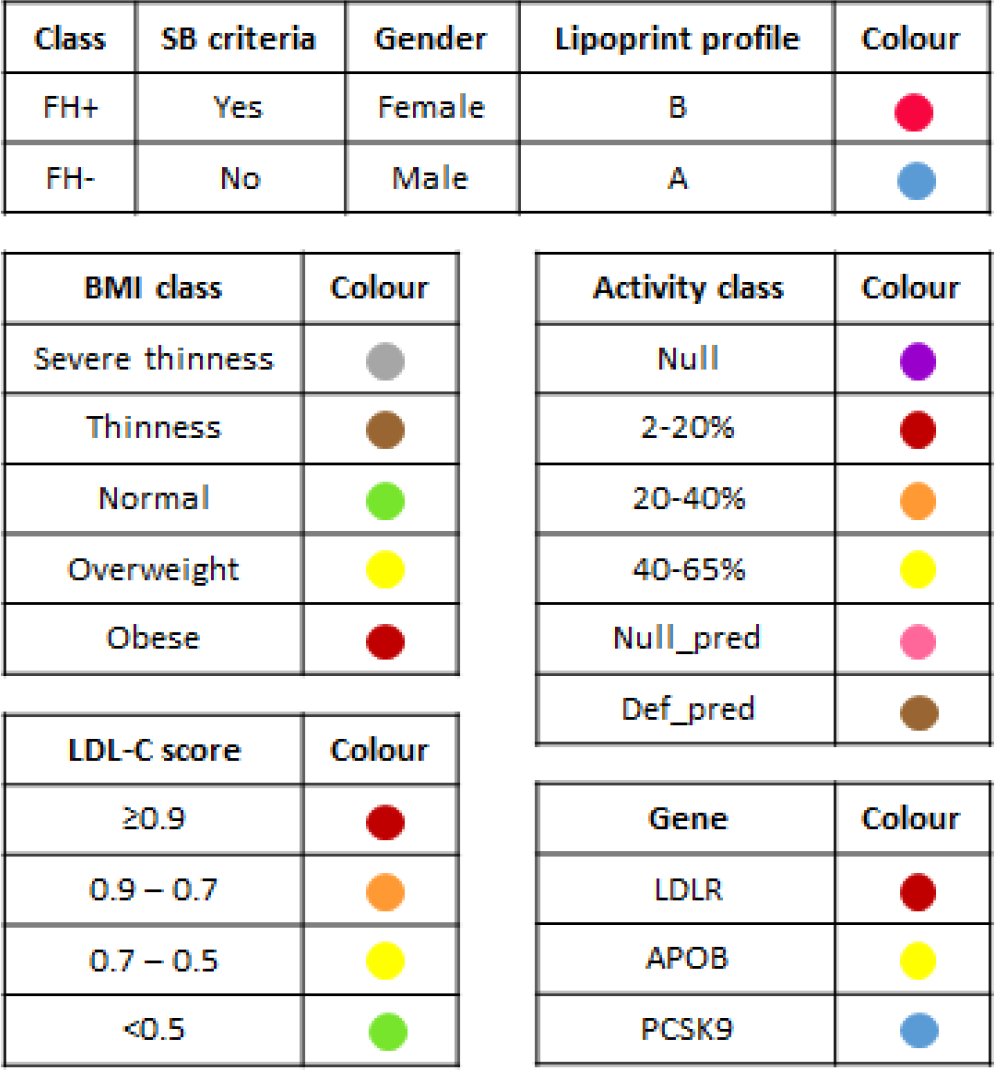
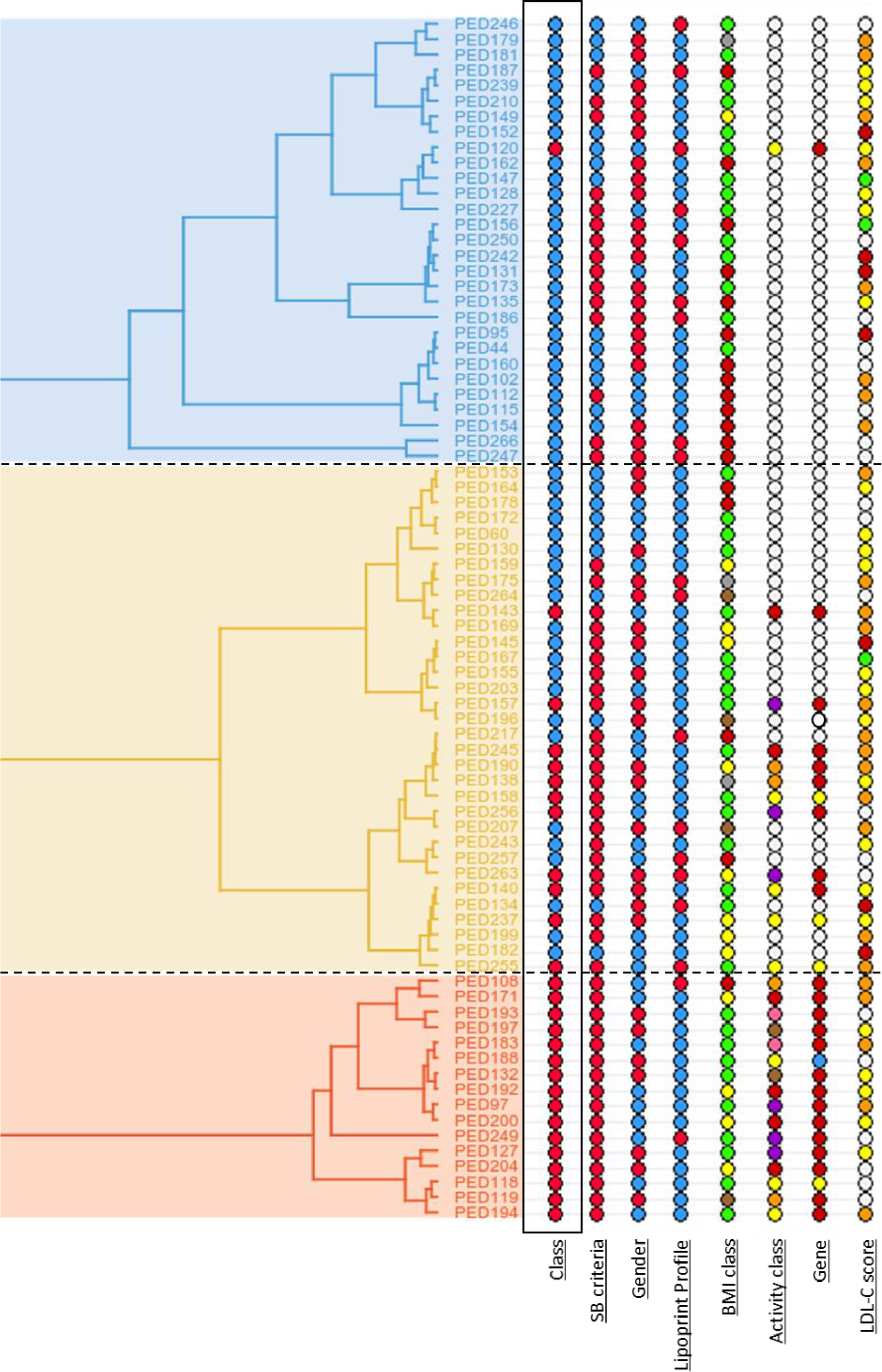
Distribution of categorical variables within the classification dendrogram. White circles on “Activity class” and “Gene” correspond to FH- individuals, while on “LDL-C score” they represent the individuals whose score was not performed.

Apart from the already recognized pattern of variable “Class”, more subtle variable distribuition patterns could be identified after careful inspection of Figure 5. Still, the “mixed cluster” could not be clearly associated with any specific combination of variables.

The most severe genetic variants corresponding to lower percentages of molecular activity, comparatively to the wild type allele, seems to predominate in the FH+ cluster, with one of the branches of the mixed cluster, presenting a higher number of individuals with intermediate alleles. This could indicate a progressive decrease in disease severity. Still, there are several individuals in the mixed cluster (i.e., PED263, PED256, PED245, PED157, PED143) that present extremely low levels of activity in the affected allele. Considering that more severe variants translate in higher levels of TC and LDL-C, other lipid parameters that were not possible to identify may be responsible for setting these FH+ individuals closer to a FH- profile, which could explain their assignment to the mixed cluster. PED120 is the only FH+ belonging to the predominant FH- cluster, which could be due to the presence of a pathogenic variant of mild effect (c.1216C>T, p.Arg406Trp), which could translate into a milder phenotype in comparison to all the other FH+ individuals [35].

The predominant FH+ cluster is totally composed of individuals that fulfil TC and LDL-C cut-offs from SB criteria, which emphasises the idea that this cluster presents the most severe phenotypes. Accordingly, the number of individuals fulfilling these cut-offs decreases as we move through the mixed cluster to the predominant FH- cluster. This implies that subjects from predominant FH+ cluster present higher levels of TC and LDL-C in comparison with individuals in other clusters. In contrast, the LDL-C score appears to be higher in the mixed cluster and, especially, within the predominant FH- cluster. This means that this last cluster presents a higher number of individuals having a stronger polygenic contribution for their biochemical profiles, namely LDL-C levels [36].

Regarding gender, there is a higher number of females in the predominant FH- cluster. The same trend is also observed for the category “obese” of the variable “BMI class”, which suggests that within the PFHS-ped dataset a FH- profile is more frequently found among girls suffering from obesity. This finding was also observed while testing the statistical association between clusters and each of the categorical variables (Figure 4). Still, gender did not show a significant association with the cluster partition (Table 3) and the predominant FH- cluster was the only cluster that presented an unbalanced gender distribution.

As mentioned before, the distribution pattern of “Gene” variable is in agreement with the classification of individuals as FH+ or FH- and, since *LDLR* variants are identified in a big majority of FH cases, this explains the considerable presence of *LDLR* variants in the predominant FH+ cluster and, in less extent, within the mixed cluster. In addition, there are four *APOB* variants among these two clusters, three of them in the mixed cluster. *APOB* variants are known to produce milder phenotypes in comparison with *LDLR* variants [9]. This highlights the differences in disease severity between clusters.

In relation to “Lipoprint profile”, the predominant FH+ cluster seems to present less individuals of profile B in relation to the other clusters. As explained before (see methods), the Lipoprint profile is obtained by measuring the concentration of sdLDL particles in serum, which correspond to the most atherogenic fraction of LDL. As such, a type B profile is related to a higher cardiovascular risk, since a higher concentration of these particles are present in comparison to a subject with a type A profile [37], [38]. This observation could be hinting at the potential influence of altered hepatic lipase (HL) activity and consequent change in the proportions of LDL subfractions on disease severity, as explained in the discussion.

### Cluster description by individuals emphasises the presence of different dyslipidaemic profiles besides the categorization as FH+ and FH-

In addition to the statistical analysis that tested the variable contribution for cluster definition, we further assessed which individuals best describe, and/or are more specific of each cluster, by measuring the distance between each individual and the gravity centre of each cluster. Table 4 presents the paragons, which are the individuals considered to best characterise each of the clusters due to their proximity the to the cluster centre.

**Table 4.**
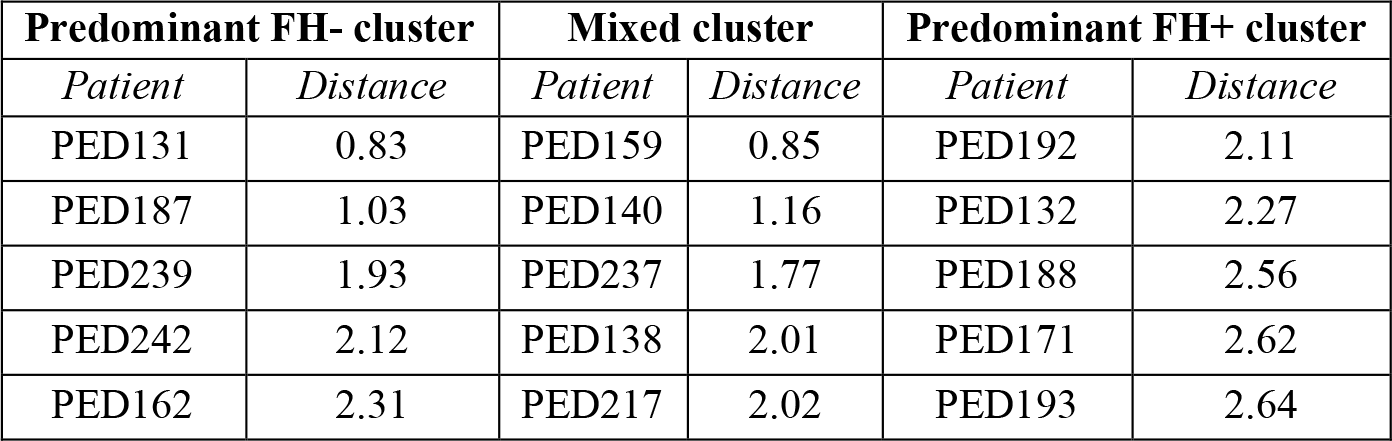
Individuals with the shortest distance to the centre of the cluster they belong.

| **Predominant FH- cluster** |  | **Mixed cluster** |  | **Predominant FH+ cluster** |  |
| --- | --- | --- | --- | --- | --- |
| *Patient* | *Distance* | *Patient* | *Distance* | *Patient* | *Distance* |
| PED131 | 0.83 | PED159 | 0.85 | PED192 | 2.11 |
| PED187 | 1.03 | PED140 | 1.16 | PED132 | 2.27 |
| PED239 | 1.93 | PED237 | 1.77 | PED188 | 2.56 |
| PED242 | 2.12 | PED138 | 2.01 | PED171 | 2.62 |
| PED162 | 2.31 | PED217 | 2.02 | PED193 | 2.64 |

As shown in Table 4, PED131, PED159 and PED192 are the individuals that best characterise predominant FH-, mixed and predominant FH+ cluster, respectively. From these three subjects, only PED192 is FH+, while PED131 is the one presenting the highest BMI and polygenic score. This is in agreement with previous results (Table 3 and Figure 4) that suggested a more severe dyslipidaemic profile for patients of predominant FH+ cluster, while a higher polygenic contribution and BMI might be associated with the predominant FH- cluster. Furthermore, the individuals that best represent the predominant FH+ cluster are mostly carriers of more severe FH-associated gene variants, with a normal BMI and a moderate polygenic contribution. On the contrary, predominant FH- cluster are best described by FH- individuals that in average present a high polygenic score and a higher BMI than normally expected, with some of them also presenting milder TC and LDL-C levels in comparison to individuals from other clusters. The mixed cluster appears to represent a mixed phenotype with characteristics of both FH+ and FH- profiles, for the following reasons: there is only one FH+ individual among the list of representative individuals. It is noteworthy that this individual presents a pathogenic variant in the *APOB* gene is considered milder than the *LDLR* variants associated to the predominant FH+ cluster [9]. The pattern of polygenic contribution in these individuals seems to be similar to those of the predominant FH+ cluster, although the polygenic scores of PED188 and PED193 were not available. Finally, the BMI pattern is worse in comparison to the predominant FH+ cluster but milder than the one found in the predominant FH- cluster; TC and LDL-C levels are higher than those of the predominant FH- cluster. Still, the low number of individuals involved in this analysis (five individuals as best representatives of each cluster) should be taken into consideration for further discussion around the mixed population.

As counterpoint to this analysis, Table 5 presents the individuals that are more distant to the centre of other clusters and that can be considered the most specific individuals of their own cluster.

**Table 5.**
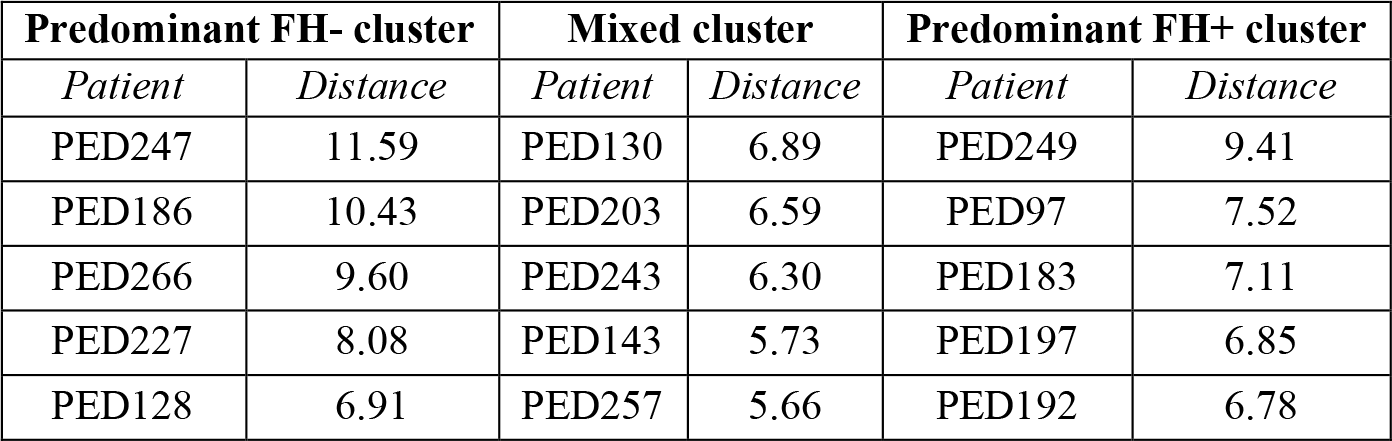
Individuals with the longest distance to the centre of other clusters, according to the cluster they belong.

As shown in Table 5, PED247, PED130 and PED249 are the most specific individuals of predominant FH-, mixed, and predominant FH+ cluster, respectively. PED249 presents a normal BMI and high levels of TC and LDL-C, besides carrying a null variant in the *LDLR* gene, which is in agreement with the pattern already associated with the predominant FH+ cluster. Indeed, the most specific individuals of this cluster have mostly high values for TC and LDL-C and present in average a moderate to high polygenic contribution and a normal BMI. Additionally, three of them are carriers of more severe gene variants. The most specific individuals of the predominant FH- cluster mostly present TC and LDL-C levels that fulfil the cut-offs from SB criteria, and a pro-atherogenic profile regarding the concentration of sdLDL measured by Lipoprint assay. In addition, two of these individuals were considered obese. Regarding the mixed cluster, the most specific individuals mostly present TC and LDL-C levels above SB cut-offs, besides of a normal BMI and a moderate polygenic contribution. PED143 is the only FH+ individual of the mixed cluster, with a gene variant associated with 2-20% of molecular activity in the affected allele, which is accompanied by a very high polygenic score. Still in the same cluster, PED257 is the only patient with a high BMI and a pro-atherogenic Lipoprint profile. As mentioned before, we should be careful while taking conclusions from these results, considering the low number of individuals involved.

PED192 belongs simultaneously to the lists of Table 4 and Table 5 regarding the predominant FH+ cluster, which means that this individual is not only among those who best characterise this cluster but also one of the most specific of its members. This patient presents high levels of LDL-C and TC, a high ratio apoB/apoA-I, besides carrying a pathogenic variant associated with 2-20% of molecular activity in the affected allele. This pattern is aligned with previous findings mentioned in this results section, regarding the predominant FH+ cluster.

### Cluster characterization by dimensions highlights the presence of different metabolic patterns among individuals

To acquire a more detailed cluster description, in addition to the characterization by variables and individuals, we assessed the PCA dimensions that best explain the distribution of individuals among clusters. For this purpose, the association between individual coordinates within each cluster and the different dimensions (also known as principal components or axes in the context of PCA results) were tested, identifying the dimensions that best describe each cluster. Table 6 shows the dimensions where individuals present the statistically significant stronger or weaker coordinates, for each of the three clusters.

**Table 6.** Dimensions that present statistically significant associations with individual coordinates within each cluster, under the confidence level of 95%.

| **Predominant FH- cluster** |  |  |
| --- | --- | --- |
| *Dimensions* | *p-value* | *Association* |
| PC2 | 5.22E-08 | stronger |
| PC4 | 0.005896 | stronger |
| PC1 | 0.000114 | weaker |
| **Mixed cluster** |  |  |
| *Dimensions* | *p-value* | *Association* |
| PC3 | 0.0394 | weaker |
| PC4 | 2.17E-05 | weaker |
| PC2 | 1.79E-06 | weaker |
| **Predominant FH+ cluster** |  |  |
| *Dimensions* | *p-value* | *Association* |
| PC1 | 7.11E-12 | stronger |

As depicted in Table 6, individuals from the predominant FH- cluster have a stronger association with PC2 and PC4 in comparison to other clusters, while this association is weaker for PC1. Conversely, individuals from the predominant FH+ cluster present a stronger association with PC1. Concerning the mixed cluster, these individuals have a weaker association with PC3, PC4 and PC2 in relation to individuals of other clusters.

Analysing the PCA results, it is possible to access which variables most contribute to each dimension. Crossing this information with the dimensions that are significantly associated with each cluster (Table 6) allowed us to consolidate the description of clusters. Given the fact that only the first five dimensions were used for the HCPC analysis (see methods), and from these only four presented a significant correlation between at least one of the three clusters (Table 6), the variable contribution by dimension was analysed only for PC1, PC2, PC3 and PC4 (Figure 6).

**Figure 6.**
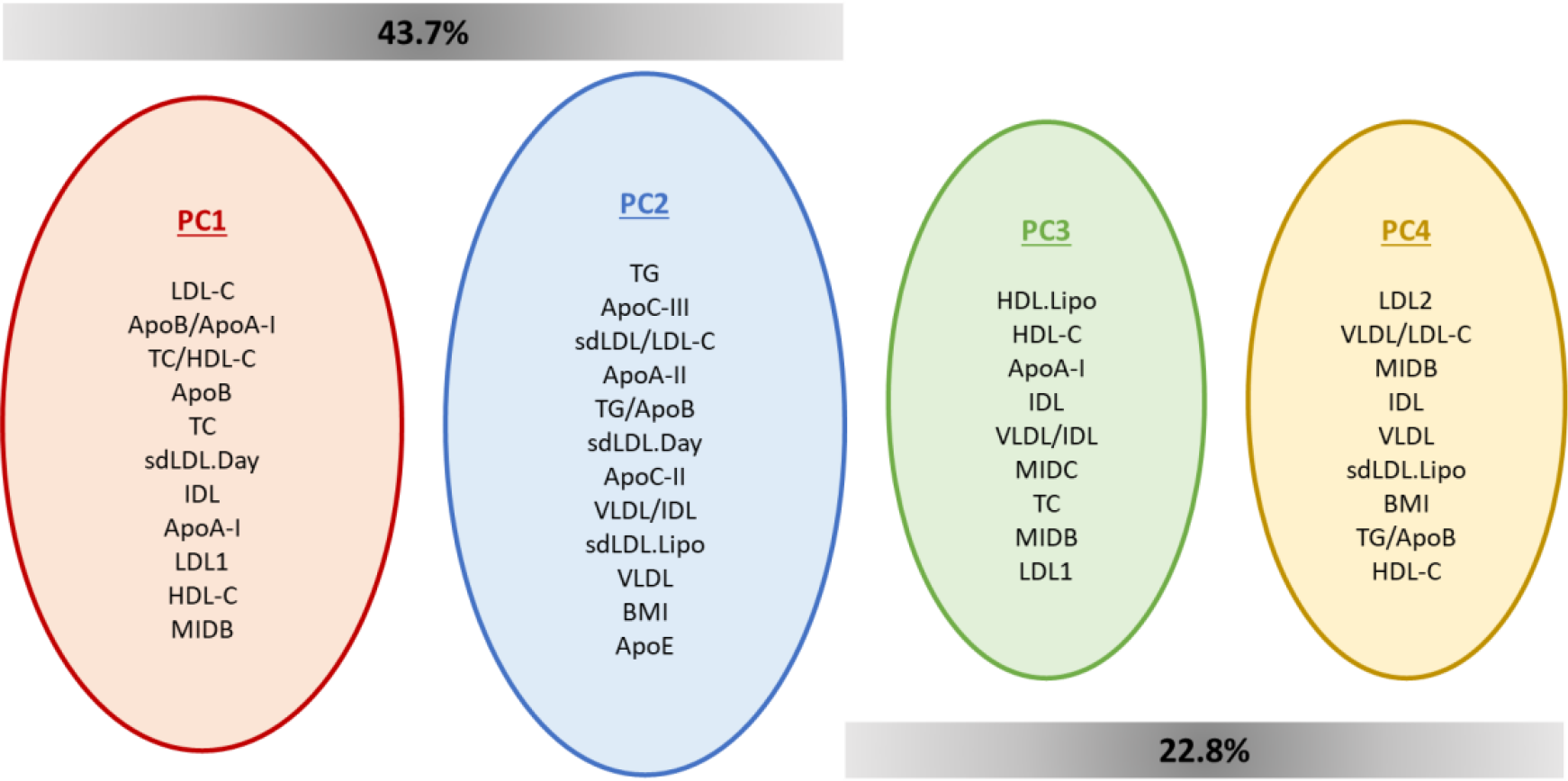
Variables that most contribute for each of the dimensions that previously have shown to be significantly correlated to clusters. PC1 and PC2 together contribute to explain 43.7% of variance in the dataset, while PC3 and PC4 together explain 22.8% of data variance.

The variables that contribute most for PC1 were mainly linked to LDL/apoB metabolism, while PC2 were mostly associated with variables related to TG metabolism and ratios that establish interactions between TG and LDL/apoB pathways. This emphasises the relation that was already established between the predominant FH+ cluster (showing a stronger association to PC1) and LDL/apoB metabolism. Conversely, the predominant FH- cluster, presenting a weaker association to PC1 and a stronger association to PC2, was previously associated with TG metabolism. This cluster also presented a strong correlation with PC4, whose main contribution came from variables associated to the different LDL fractions, besides other parameters like BMI, TG/apoB and HDL-C. In contrast, PC4 presented a weaker association with the mixed cluster, which was also reported for PC2 and PC3, related to parameters involved in TG metabolism and LDL pathway/reverse cholesterol transport, respectively. This translates to lower values of these parameters for individuals from the mixed cluster in comparison to individuals from other clusters.

Given the fact that PC1 and PC2 explain more than 40% of the data variance, which makes them the most informative dimensions, the distribution of individuals within each cluster was plotted for PC1 and PC2 dimensions (Figure 7).

**Figure 7.**
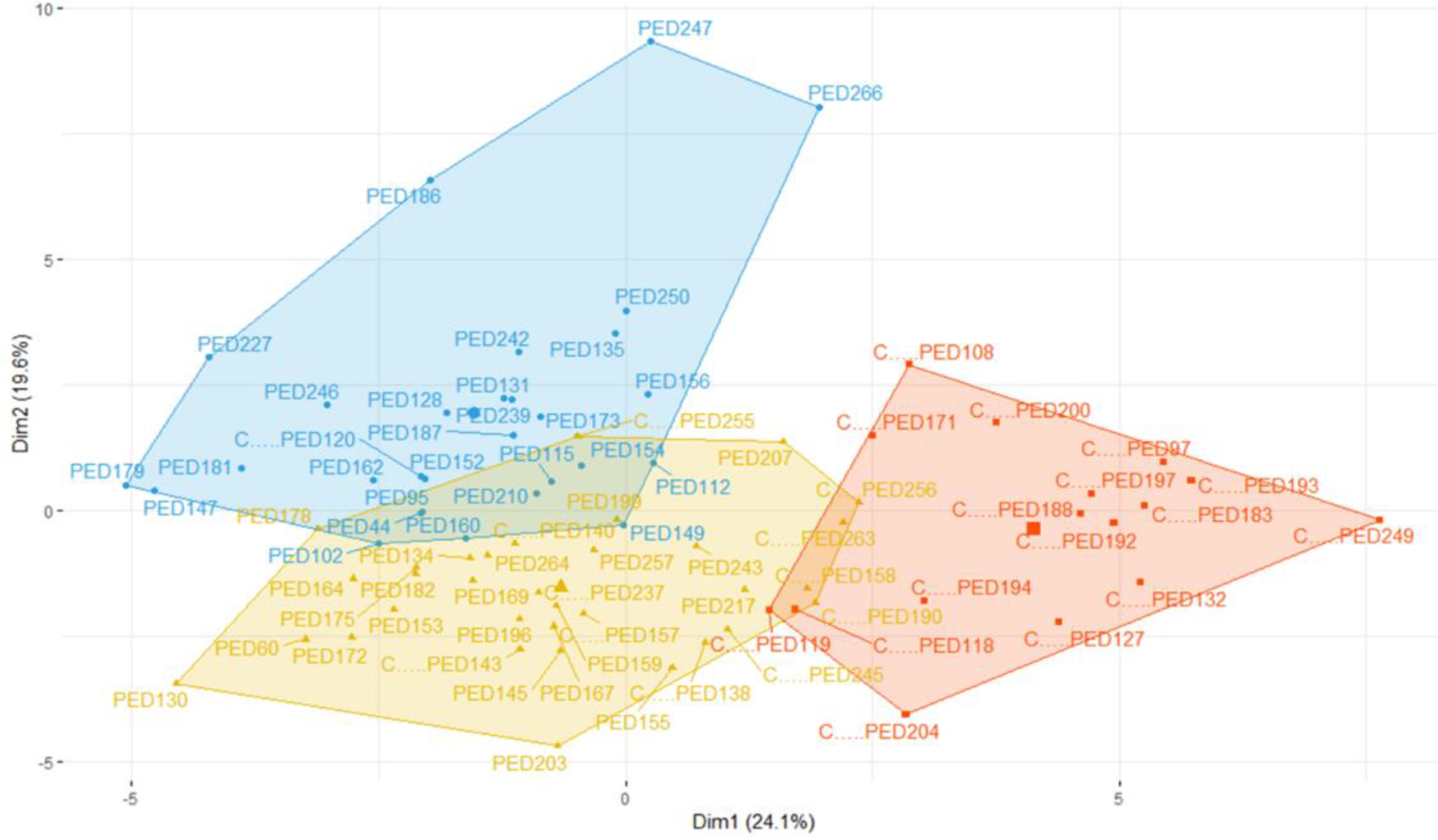
Plot showing the distribution of individuals within each cluster for PC1 (also known as Dim1) and PC2 (also known as Dim2), which correspond to the dimensions that best explain data variance. FH+ individuals are identified with a ‘C’ before their ID. Individuals belonging to the predominant FH- cluster is represented in blue, mixed cluster in yellow, and predominant FH+ cluster in red.

Figure 7 clearly shows the strong correlation of the predominant FH+ cluster with PC1, since the coordinates of individuals are all positive for this dimension. Conversely, most of the individual coordinates within the predominant FH- cluster are positive for PC2 and negative for PC1. Regarding the mixed cluster, a considerable number of individuals presents negative coordinates for both PC1 and PC2 dimensions.

### Predicted class assignment using the Imp_B FH classification model suggests the presence of borderline individuals

As explained before, contrary to the predominant FH+ and predominant FH- clusters, the mixed cluster could not be clearly characterised due to the absence of specific biochemical or clinical patterns. To try to better understand the major patterns associated to this cluster, we applied one of the FH prediction models previously developed by us (Imp_B) [12], which includes the parameters LDL-C, apoB/apoA1 and TG/apoB, to the 78 individuals of the working population to determine the associated probability of each individual belonging to the FH+ and FH- class. The difference between the probability of being FH+ and the probability of being FH- (named as Δprob) was then calculated for each individual. For a better visualisation and results interpretation, the set of values of Δprob were grouped into four categories: very ambiguous (Δprob <0.25), ambiguous (0.25 ≥ Δprob < 0.5), reasonable (0.5 ≥ Δprob < 0.75), clear (Δprob ≥ 0.75). Accordingly, a high Δprob corresponds to a clear classification, while a low Δprob is associated with an ambiguous classification. Figure 8 shows the distribution of Δprob among individuals within clusters. The predicted class assignment, associated probabilities, Δprob and correspondent categories are available for all individuals of the work population in Supplementary Table 3.

**Figure 8.**
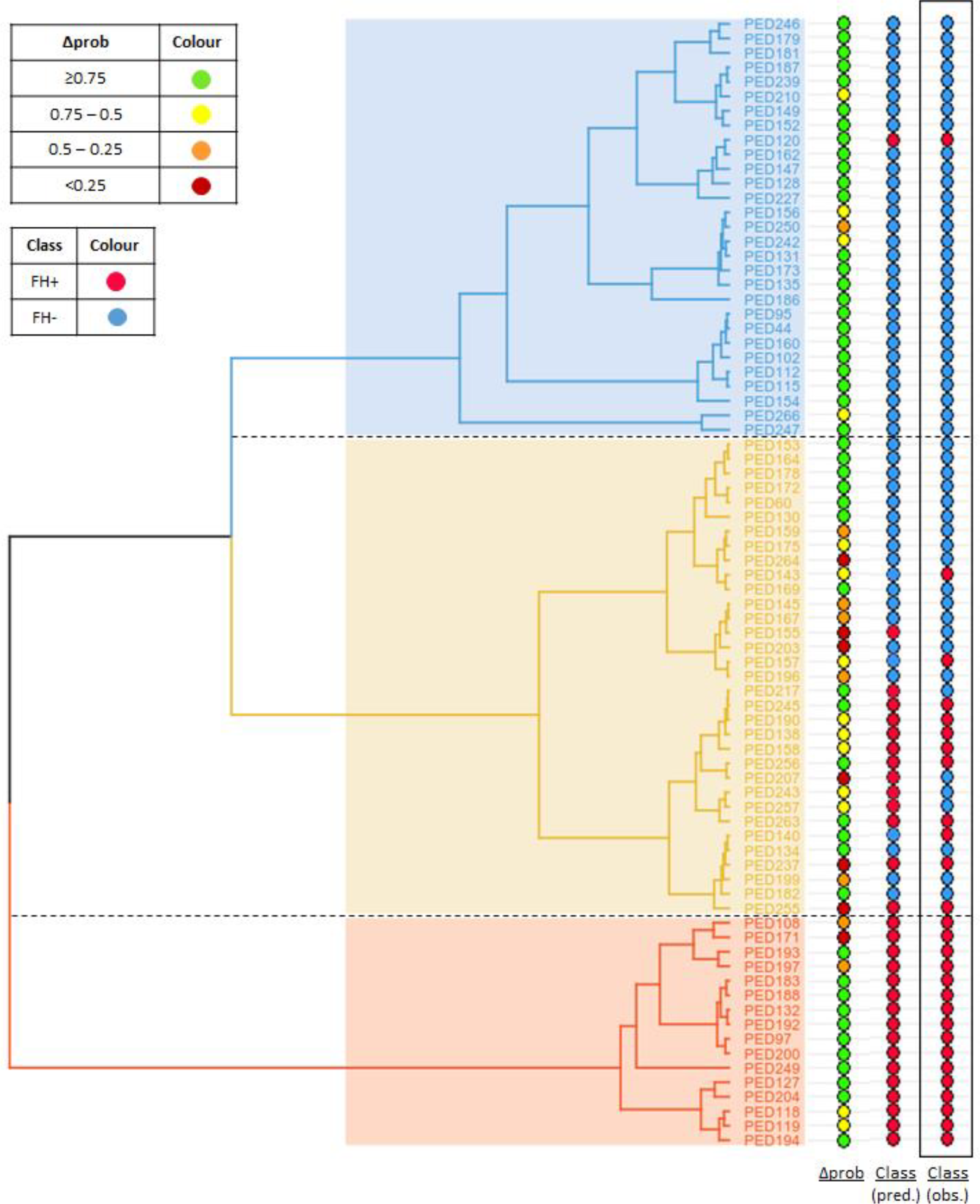
Distribution of Δprob across the 78 individuals of the work population, ordered according to cluster position. The value of Δprob is the difference between the probability of being classified as FH+ and the probability of being classified as FH- according to the predictions obtained using the Imp_B model. Both predicted and observed classifications are also shown in the figure. The colour label corresponding to the different categories of Δprob and classification (FH+/FH-) are present in the upper left corner.

As seen in Figure 8, most individuals considered to have an ambiguous classification, according to the predictions of the “Imp_B” model, are present in the mixed cluster. This is in agreement with the hypothesis that this cluster contains individuals with an intermediate biochemical phenotype that can be made evident through the application of a prediction model that considers only four parameters: LDL-C, TG, apoA1 and apoB. Thus, the FH+ individuals of this cluster seem to present a biochemical profile milder than individuals of the predominant FH+ cluster, while the FH- subjects appear to have a more severe profile than individuals of the predominant FH- cluster. Given the lack of other variables that clearly correlate with this cluster, namely, the presence of a low polygenic score, it is tempting to speculate if the FH- individuals may present a mutation in a new disease gene, or the FH+ individuals may present a disease modifying variant. Future studies of these individuals, namely through whole genome sequencing, might provide important new insights into the genetic basis of FH.

## DISCUSSION

The present study is an interesting example of the considerable potential of novel methodologies associated to data mining to contribute to a better understanding of patient classification and support the identification of new biomarkers for clinical purposes. Indeed, the unsupervised clustering approach used was able to identify distinct lipid profiles among individuals, beyond the standard genetic FH+/FH- classification. For every data subset studied, clustering analysis revealed the presence of three group of patients presenting a distinct pattern regarding the prevalence of FH+ and FH- individuals. In addition to two clusters mainly composed of FH+ and FH- individuals, a third cluster defined a mixed population with a considerable number of both genetic traits. The differences among clusters were best defined in the data subset that incorporated the highest number of biochemical parameters, emphasising the importance of using an extended lipid profile to improve the distinction between individuals.

The analysis of the quantitative variables that most contribute to the definition each cluster allowed the identification of pathways of interest in the context of lipid metabolism. Hence, the results obtained point to an important role of the LDL/apoB pathway and TG metabolism as main contributors for the lipid profiles of the predominant FH+ and the predominant FH- clusters, respectively. The association of these pathways to each group of individuals (FH+ and FH-) was already established in a previous study analysing the PFHS dataset [6]. The potential contribution of a disturbed TG metabolism to the dyslipidaemic profile of FH- individuals may be due to the influence of environmental factors, such as cholesterol and TG-rich diets [6]. Of note, the considerable presence of parameters associated to LDL subfractions (result of the VLDL – IDL – LDL delipidation cascade) among the variables that best described the predominant FH+ cluster, highlights once again the importance of an extended lipid profile to achieve a better characterisation of the individuals.

The description of clusters by categorical variables gave us additional information that supported the identification of molecular, biochemical, and anthropomorphic patterns among clusters, including suggestions for a better understanding of the distribution of individuals within the mixed cluster, where no clear pattern was detected so far. The predominant FH- cluster was found to be characterised by a significant presence of girls suffering from obesity with lower TC and LDL-C levels, in comparison to FH+ patients. In contrast, individuals of the predominant FH+ cluster presented higher levels of TC and LDL-C and severe *LDLR* pathogenic variants. Interestingly, the polygenic LDL-C score was higher in the predominant FH- cluster. These evidences suggest a considerable contribution of polygenic and lifestyle factors for the development of dyslipidaemia in FH- individuals, as hypothesised by other studies [6], [36], [40]. The use of BMI class as a supplementary variable emphasised the relevance of BMI to this phenotype as one of the quantitative variables that presented a significant contribution to cluster partition. Given the fact that BMI class was based on WHO percentiles dependent of age and sex, it takes into consideration the different phases of development during childhood and adolescence. Regarding gender, we should take into account that despite its apparent association with the predominant FH- cluster, this variable did not present a significant association with the cluster partition. Future studies using a bigger number of individuals than PFHS-ped and enrolling also normolipidaemic individuals, aiming for a better representation of the general population, would be essential to validate gender as a discriminant factor of lipid levels.

In relation to the mixed cluster, these individuals appeared to represent an intermediate phenotype, since their profile is milder than those from the predominant FH+ cluster but more severe than those from the predominant FH- cluster. Indeed, taking the dendrogram as reference, the degree of dyslipidaemia severity (mainly regarding levels of TC and LDL-C and results of the molecular study) appeared to be gradually increased as one moves towards the predominant FH+ cluster from, while the polygenic contribution appeared to gradually increase towards the predominant FH- cluster.

Although the Lipoprint assay allows us to acquire useful information regarding LDL subfractions, we should remember that it is a semiquantitative method and that the obtained measurements may not be as accurate as those achieved using quantitative methods (e.g., photometric test used for measuring parameters of “Advanced” profile) [37]. Indeed, FH+ patients present higher sdLDL concentrations when using a “daytona” assay (RX daytona+® analyser), in comparison to FH- individuals, which is expected regarding the role of sdLDL as pro-atherogenic particles within the context of FH. Conversely, Lipoprint results regarding the predominance of profile A among FH+ population and profile B within the predominant FH- cluster can be explained by an altered HL activity in FH+ patients that results in higher prevalence of LDL1/LDL2 subfractions. Alternatively, this could reflect a higher influence of lifestyle (e.g., TG-rich diets) and polygenic factors in FH- individuals [41]–[44]. Of note, although FH- subjects present lower TC and LDL-C levels in comparison to FH+ patients, they are not normolipidaemic and usually tend to present borderline values of TC and LDL-C when they fail to fulfil the cut-offs of SB criteria. This can be explained by a likely presence of a polygenic form of dyslipidaemia, resulting from the cumulative burden of several SNPs able to raise LDL-C levels [36], [40].

The characterization of clusters regarding the contribution of each dimension used for HCPC analysis highlighted the influence of different lipid patterns in individual distribution. PC1 and PC2, considered the most informative components, were associated with the previous findings regarding the connection of LDL/apoB pathway and TG metabolism with predominant FH+ and FH- clusters, respectively. Although with a smaller level of contribution, PC4 revealed a strong association with the predominant FH- cluster, while their main contributors were parameters related to LDL subfractions and others like BMI and TG/apoB. The association between this cluster and delipidation cascade might be explained by the predominance of a Lipoprint profile B in FH- individuals, as discussed above. In addition, the association of these individuals with parameters as BMI and TG/apoB emphasises the potential impact of a perturbed TG metabolism and lifestyle in the development of their dyslipidaemia. Concerning the mixed cluster, there was a negative association with PC2, PC3 and PC4. These findings might be explained by the presence of some FH+ individuals that decrease the levels of parameters associated to TG and reverse cholesterol pathways, while the simultaneous presence of a considerable number of FH- individuals decreases the levels of parameters involved in LDL/apoB pathway. The identification of a high prevalence of “borderline individuals” using our previously developed FH classification model [12], supports the hypothesis that this cluster represents a mixed phenotype of dyslipidaemia. It is intriguing to speculate that new genetic variants – both new disease or modifier genes - may be associated to the individuals in this cluster.

In summary, the application of a detailed unbiased hierarchical clustering analysis to the PFHS-ped dataset has allowed the characterisation of specific biochemical, molecular, and anthropomorphic patterns that classify individuals, beyond their FH+ or FH- status. Future studies based on the observations presented here may contribute to the identification of potential biomarkers and pathways of interest for lipid metabolism disorders.

## Data Availability

All data produced are available online at the GitHub repository.

https://github.com/GamaPintoLab/MartaCorreia-PhD-thesis

## Acknowlegments

We are thankful to Francisco R. Pinto for discussion of the results. This work was supported by UIDB/04046/2020 and UIDP/04046/2020 Centre grants from FCT, Portugal (to BioISI). MC is recipient of a fellowship from the BioSys Ph.D. programme PD65-2012 (Ref PD/ BD/114387/2016) from FCT (Portugal).

## Author contributions

M.C. implemented the analysis, produced and discussed results, prepared all manuscript figures and data, and wrote the manuscript; M.B. is responsible for the PFHS and the production of the respective biochemical and genetic data, she defined the problem to be addressed in this study and contributed to the discussion of the results; M.G.C. defined the research approach, supervised the research work, analysed and interpreted the results, and wrote the manuscript.

## Data availability

Data used in this study are available through reference [12] and as Supplementary file 1.

