## Supplementary Figure 1 for "Analysis of a paediatric cohort of dyslipidaemic patients using unsupervised learning methods provides insights into the biochemical phenotypes of familial hypercholesterolemia"

**Supplementary Figure 1.** Distribution of parameters by clusters, with an asterisk showing the clusters whose mean was significantly different from the overall mean, under a confidence level of 95%. Parameters were grouped according to the mean trend among clusters, as present in the panel label. Clusters are identified according to the cluster label.

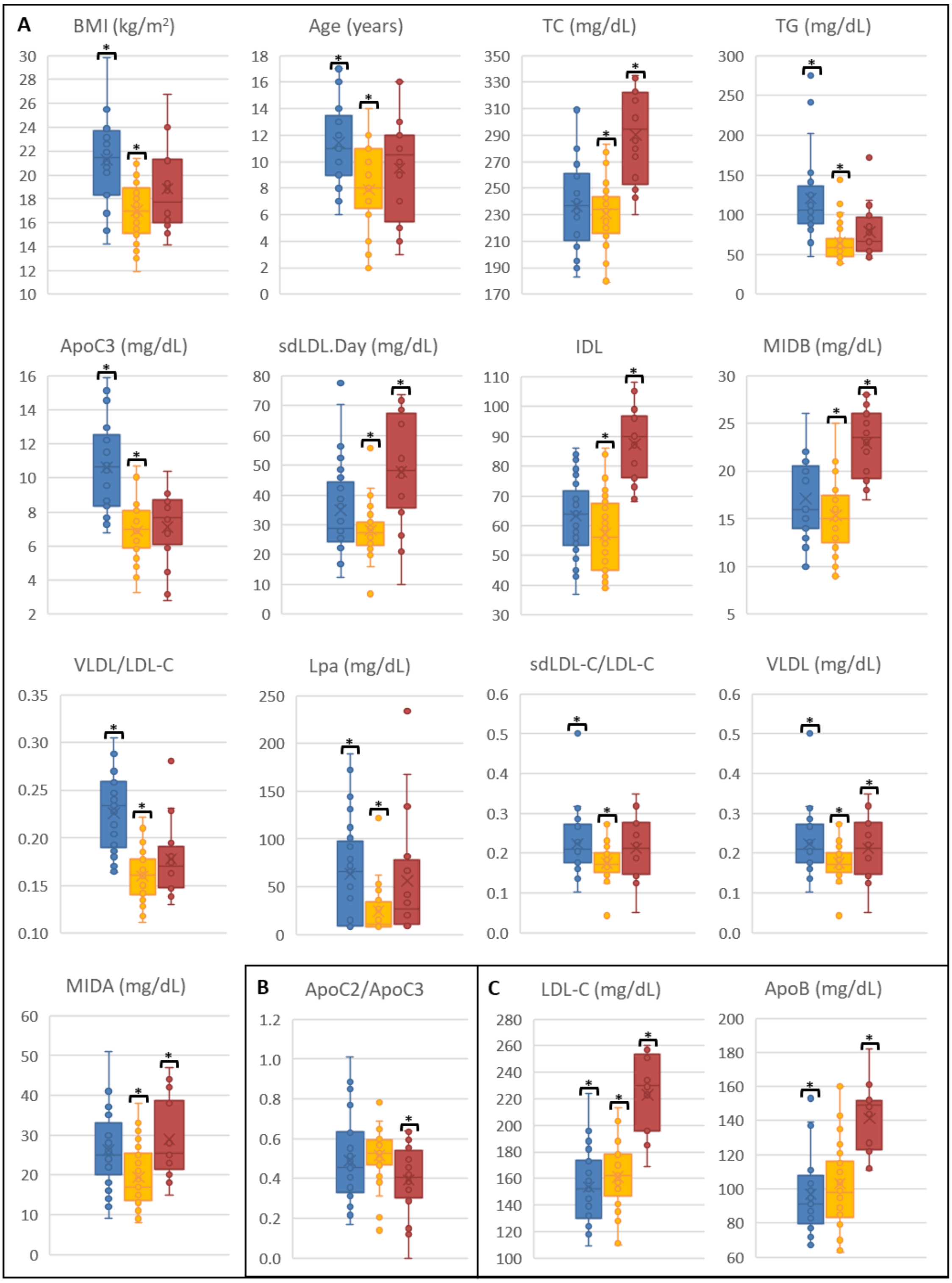

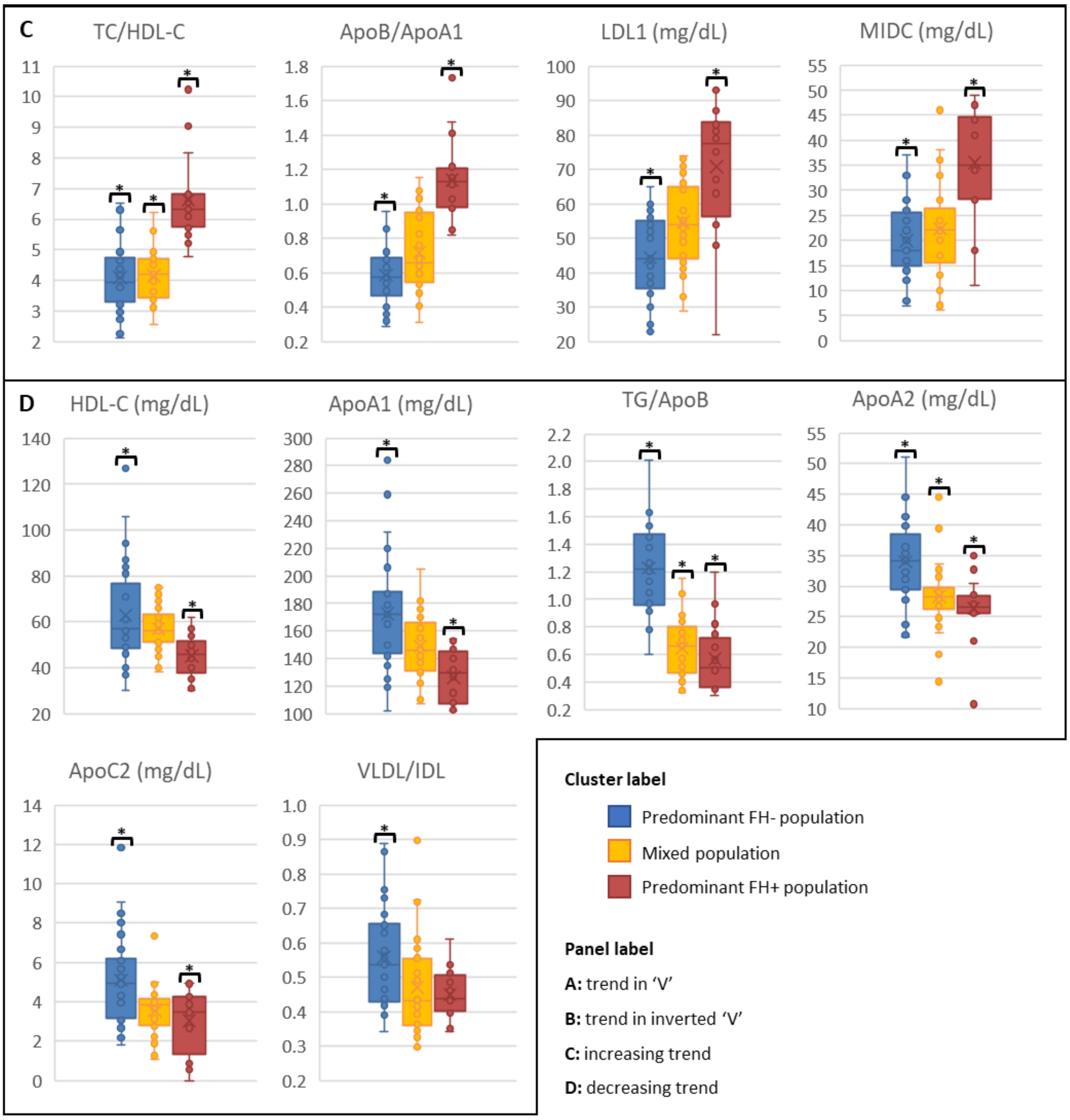
C
TC/HDL-C
ApoB/ApoA1
LDL1 (mg/dL)
MIDC (mg/dL)
D
HDL-C (mg/dL)
ApoA1 (mg/dL)
TG/ApoB
ApoA2 (mg/dL)
ApoC2 (mg/dL)
VLDL/IDL
Cluster label
Predominant FH- population
Mixed population
Predominant FH+ population
Panel label
A: trend in 'V'
B: trend in inverted 'V'
C: increasing trend
D: decreasing trend
